# Trends in the Proportion of Women as peer reviewers in JAMA, 2009-2018

**DOI:** 10.1101/2020.05.12.20088419

**Authors:** Lisa Kipersztok, Gwinyai Masukume, Julio González-Álvarez, Victor Grech

## Abstract

The achievement of gender equity by 2030 is one of the international Sustainable Development Goals adopted by United Nations member states. Peer review is crucial to academia and diverse perspectives add significant value by avoiding publication biases. We investigated the trend in female peer reviewers in *JAMA*, a globally influential medical journal, over the past decade. Based on publicly available data with a sample size of 33,745, we found an increased proportion of female peer reviewers from 23.9% in 2009, to a peak of 29.1% in 2018. Despite an increase in the proportion of female peer reviewers over the past decade, if we assume a linear trend, gender equity in peer reviewers for *JAMA* would not be reached until 2065, beyond the 2030 Sustainable Development Goal target.

## Introduction

The achievement of gender equity by 2030 is one of the international Sustainable Development Goals adopted by United Nations member states, an avowed means to improve health and wellbeing for all populations.^1^ Although women remain underrepresented in academic medicine, their representation has improved over the past decade.^2^ Just one decade ago, the proportion of female peer reviewers was less than 30% for *JAMA* and five other leading international general medical journals.^3^ Peer review is crucial to academia and diverse perspectives add significant value by avoiding publication biases.^4^ We investigated the trend in female peer reviewers in *JAMA*, a globally influential medical journal, over the past decade.

## Methods

We used publicly available data on *JAMA* peer reviewers from 2009 through 2018.^5^ Institutional review board approval was not required because of the public nature of this dataset. We determined reviewer gender by way of their first name, using https://gender-api.com/, which has an extensive database of forenames linked to gender. Given our sample size of 33,745, assuming a 5% error margin and a 95% confidence interval, we randomly selected 400 reviewers for manual gender identification through online searches for pronouns like he/she and photographic information.

## Results

From the random sample of 400, after several attempts, the gender of nine peer reviewers were unable to be determined. The remaining 391 reviewers corresponded to 274 males (70.1%) and 117 females (29.9%). The agreement between manual and automatic gender classification was statistically adequate at 96.9%.

The proportion of female reviewers increased over the study period, from 23.9% in 2009, to a peak of 29.1% in 2018 as depicted in the **Figure**. Assuming a linear trend, the percentage of women reviewers increased by 0.4% annually (Spearman’s rho, p = 0.008) over the decade.

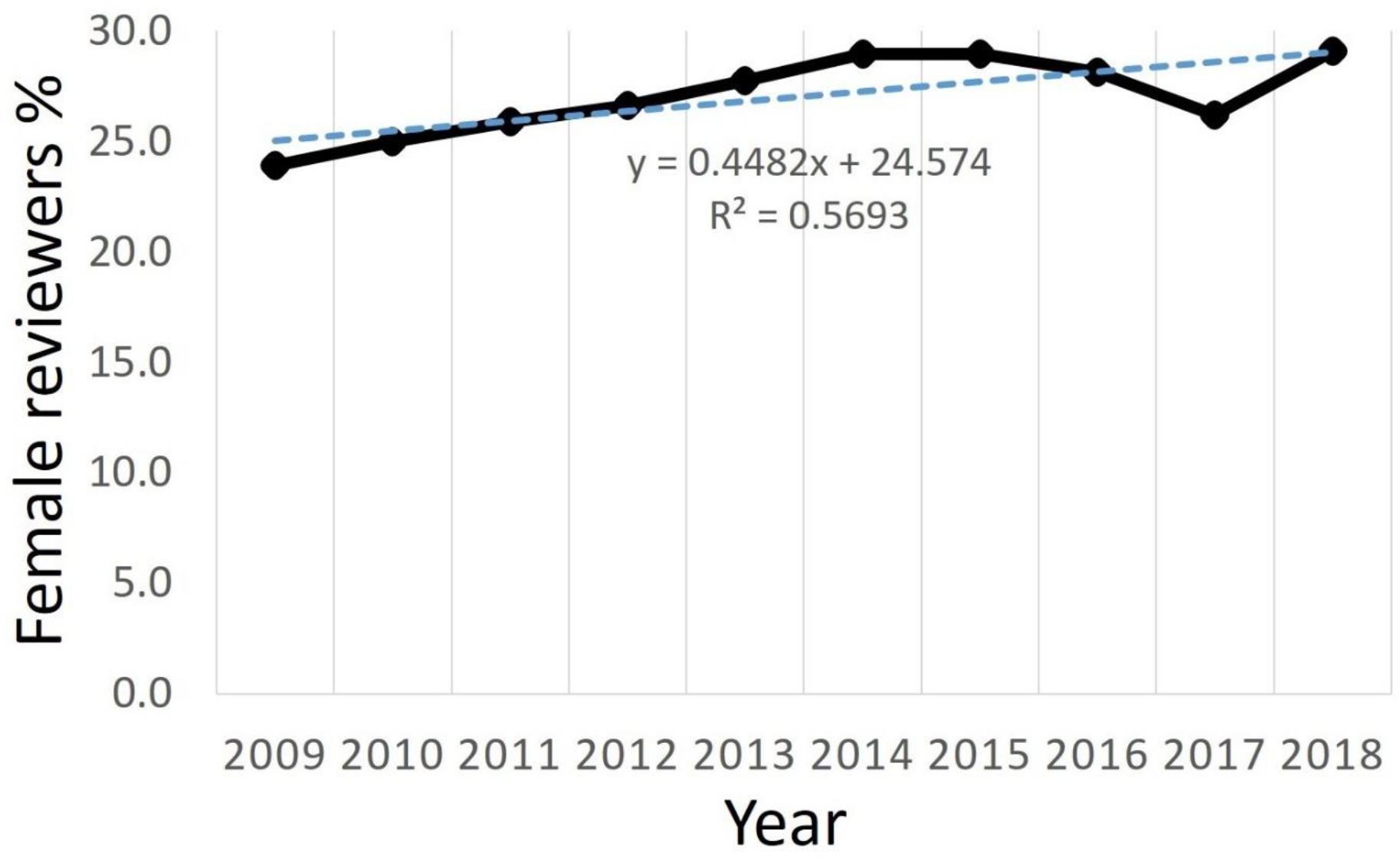
**Figure**. Temporal trends for female peer reviewers.

## Discussion

We found an increased proportion of female peer reviewers from 2009 to 2018. However, if the current trend is maintained, in 2030, only 34% of *JAMA* peer reviewers will be females. Assuming the same trend, gender equity in peer reviewers for *JAMA* would not be reached until 2065.

The limitations of this study include the assessment of one journal. Additionally, we were unable to determine non-binary representation using our gender assessment method. We suspect that the trend and proportion of female peer reviewers would be similar in other international and local medical journals, given the global underrepresentation of females in academic medicine.^1, 3^ Given the equivalent enrolment of females and males in United States medical schools and other jurisdictions for over two decades,^6^ the low numbers of female peer reviewers persisting over the last decade (albeit with some improvement), is heartening but troubling. Given that peer reviewers tend to be older physicians, there may be a substantial lag period between increasing female physicians in academia and reaching equity. Continuing to measure and quantify female representation in academic medicine is a key step in achieving gender equity.^1, 6^

## Data Availability

Publicly available data was used.

